## Supplementary Text and Tables for "A mechanistic and data-driven reconstruction of the time-varying reproduction number: Application to the COVID-19 epidemic"

### Supporting Information

#### State-space framework

We have used a state-space framework defined by two sets of equations: the first set describes the propagation of the disease in the population and the second corresponds for the observation process. This allows to consideration of unknowns and uncertainty both in the epidemiological process and in the observation of epidemic dynamics:

$$\begin{cases} \dot{x}(t) = g\left(t, x(t), \theta(t), u(t)\right) \\ y(t) \mid x(t) \sim f\left(h(x(t)), \phi(t)\right) \end{cases} \qquad \text{(S1)}$$

The first equation of (S1) corresponds to the epidemiological model, with *x(t)* representing the state variables (for instance, $S(t)$ the susceptibles, $E(t)$ the infected non-infectious, $I(t)$ the infectious and $R(t)$ the removed for the classical SEIR model) and $\theta(t)$ the epidemiological parameters, among which some are time-dependent. The second equation is associated to the observational process defined by the probabilistic distribution $f$ with parameters depending on a functional of some of the epidemiological model coordinates, $h(x(t))$ and some observational and/or measurement error, $\phi(t)$. $y(t)$ corresponds to observed epidemic dynamics (partial and noisy observations of $x(t)$) and $u(t)$ is the process noise describing intrinsic or environmental stochasticity other than the observational noise is included in $f$. In our applications, $h(x(t))$ will represent the daily new cases.

***Epidemiological Model – hidden state***

Our objective was to propose a simple but relevant model that incorporates the available information to examine the dynamics of Covid-19 epidemics before, during and after the lock-down.

Our model is an extended stochastic SEIR model also accounting for asymptomatic transmission and the hospital course of the hospitalized infectious. It is similar to other models that have already been proposed to model and forecast the Covid-19 epidemic (Li et al, 2020; Prague et al, 2020, Di Domenico et al, 2020; Liu et al, 2020; Prem et al, 2020). It includes the following variables the susceptibles $S$, the infected non-infectious $E$, the symptomatic infectious $I$, the asymptomatic infectious $A$, the removed people $R$, and the hospital-related variables: hospitalized people $H$, people in intensive care unit $ICU$, cured people $G$, and deaths at hospital $D$. We have also introduced Erlang-distributed stage durations (with a shape parameter equal to 2) in $E$, $I$, $A$ and $H$ compartments, to relax the mathematically convenient but unrealistic assumption of exponential stage durations. As more and more people have been vaccinated and as the duration of the immunity conferred shows a good persistence of the antibodies induced (Widge et al, 2021), the effect of vaccination is introduced simply by considering the effect of vaccination on the depletion of susceptibles. For sake of simplicity, we present here below the differential equations describing the deterministic version of our model, whereas its stochastic version was used in this study:

$$\frac{dS}{dt} = -\beta(t).\left(I_1 + q_1.I_2 + q_2.(A_1 + A_2)\right) - V$$
$$\frac{dE_1}{dt} = \beta(t).\left(I_1 + q_1.I_2 + q_2.(A_1 + A_2)\right) - \frac{\sigma}{2}.E_1$$
$$\frac{dE_2}{dt} = \frac{\sigma}{2}.(E_1 - E_2)$$
$$\frac{dI_1}{dt} = (1-\tau_A).\frac{\sigma}{2}.E_2 - \frac{\gamma}{2}.I_1$$
$$\frac{dI_2}{dt} = \left(1 - \tau_H - q_I.\tau_I\right)\frac{\gamma}{2}.I_1 - \frac{\gamma}{2}.I_2$$
$$\frac{dA_1}{dt} = \tau_A.\frac{\sigma}{2}.E_2 - \frac{\gamma}{2}.A_1$$
$$\frac{dA_2}{dt} = \frac{\gamma}{2}.(A_1 - A_2)$$
$$\frac{dR}{dt} = \frac{\gamma}{2}.\left(\left(1 - \tau_H - q_I.\tau_I\right).I_2 + A_2\right) + (1 - q_D.\tau_D).\frac{\kappa}{2}.H_2 + V \qquad \text{(S2)}$$
$$\frac{dH_1}{dt} = \tau_H.\frac{\gamma}{2}.\left(I_1 + I_2\right) - \frac{\kappa}{2}.H_1$$
$$\frac{dH_2}{dt} = \left(1 - \tau_I - q_D.\tau_D\right).\frac{\kappa}{2}.H_1 + (1-\tau_D).\delta.ICU - \frac{\kappa}{2}.H_2$$
$$\frac{dICU}{dt} = q_I.\tau_I.\frac{\gamma}{2}.\left(I_1 + I_2\right) + \tau_I.\frac{\kappa}{2}.H_1 - \delta.ICU$$
$$\frac{dG}{dt} = \left(1 - q_D.\tau_D\right).\frac{\kappa}{2}.H_2$$
$$\frac{dD}{dt} = q_D.\tau_D.\frac{\kappa}{2}.\left(H_1 + H_2\right) + \tau_D.\delta.ICU$$

The main originality of this model is the time-varying transmission rate *β(t)* that follows a Brownian diffusion process, equation (1) in the main text. $\sigma$ is the incubation rate and $1/\sigma$ is the average incubation period. As in previous models (*e.g.* Di Domenico et al, 2020; Fergusson et al, 2020), we reduce the incubation period duration to account for pre-symptomatic infectious. In this way individual that become infectious before the end of the infectious period but remain either asymptomatic or develop symptoms are put in the infectious compartment. The other parameters are: $\gamma$ the recovery rate, $1/\kappa$ the average hospitalizion period, $1/\delta$ the average time spent in ICU, $\tau_A$ the fraction of asymptomatics, $\tau_H$ the fraction of infectious hospitalized, $\tau_I$ the fraction of ICU admission, $\tau_D$ the death rate, $q_1$ and $q_2$ the reduction in transmissibility of $I_2$ and asymptomatics ($A_1$+$A_2$), respectively. As the peaks of hospitalized and admitted to ICU individuals are concomitant, we consider that a small fraction, $q_I.\tau_I$ of infectious with severe symptoms enter directly the ICU. Even if the majority of deaths occur in the ICU, a fraction, $q_D.\tau_D$, can occur in the hospital, outside the intensive care. Then, $q_I$ and $q_D$ are the reduction in admission in ICU and in death rate, respectively. Most of the parameters are estimated in our inference process and the remaining ones set to plausible values, in agreement with the literature (see S1 Table). In this model, variable *V* represents vaccinated people and can be interpreted as "effectively protected vaccinated people". *V* is proportional to the number of individuals vaccinated with one and/or two doses as:

$$V(t) = e_1.V_{dose1}(t - d_e) + e_2.V_{dose2}(t - d_e) \quad \text{(S3)}$$

where $V_{dosei}$ is the number of individuals vaccinated with one or two doses, $d_e$ the delay after vaccination and $e_i$ is the vaccine efficiency. For this latter parameter, the range documented in the literature is between [17%-73%] for one dose and [52 %-93%] for 2 doses (Bernal et al, 2021; Dagan et al, 2021; Hall et al, 2021; Moustsen-Helms et al, 2021; Thompson et al, 2021), by considering all types of available vaccines. Of course, the vaccine efficiency depends on population analyzed, the vaccine used and the delay after vaccination. The delay before protection varies according to the studies between 1 week and 5 weeks (Bernal et al, 2021; Dagan et al, 2021; Hall et al, 2021; Moustsen-Helms et al, 2021; Thompson et al, 2021). For the AstraZeneca/Oxford only the effect of one dose has been studied in UK (Bernal et al, 2021). Taking these analyses into consideration we used conservative values: $e_1 = 0.45$; $e_2 = 0.85$ and $d_e = 14$ days.

Our inference approach is mainly based on data on new entries in several model compartments. These observed variables used are described in the following equations:

$$C_I(t) = \int_t^{t+1} (1-\tau_A).\frac{\sigma}{2}.E_2(t).dt$$

$$C_H(t) = \int_t^{t+1} \tau_H.\frac{\gamma}{2}.\left(I_1(t) + I_2(t)\right).dt$$

$$C_{ICU}(t) = \int_t^{t+1} \left(q_I.\tau_I.\frac{\gamma}{2}.\left(I_1(t) + I_2(t)\right) + \tau_I.\frac{\kappa}{2}.H_1(t)\right).dt \quad \text{(S4)}$$

$$C_G(t) = \int_t^{t+1} \left(1 - q_D.\tau_D\right).\frac{\kappa}{2}.H_2(t).dt$$

$$C_D(t) = \int_t^{t+1} \left(q_D.\tau_D.\frac{\kappa}{2}.\left(H_1(t) + H_2(t)\right) + \tau_D.\delta.ICU(t)\right).dt$$

where $C_I$ is the incidence of symptomatic cases, $C_H$ the daily admission to hospital, $C_{ICU}$ the daily new admission in ICU, $C_D$ the new daily number of of hospital deaths and $C_G$ the daily new discharge at hospital.

***Observational Model***

Our model trajectories are related to the data using a negative binomial observation model, which is commonly used (Bretó et al, 2009). The observed incidences, $C_{k,obs}(t)$, are assumed to be drawn from a negative binomial distribution with mean $\rho_k.C_k(t)$ and variance $\rho_k.C_k(t) + \phi_k.(\rho_k .C_k(t))^2$. For each $k$, $C_k(t)$ represents the number of events occurring during day $t$ simulated by the model, $\rho_k \in [0,1]$ is an observation rate (or reporting rate) and $\phi_k$ is an over-dispersion parameter.

Current hospital data, $H_{obs}(t)$ (corresponding to $H_1+H_2+ICU$) and $ICU_{obs}(t)$ (corresponding to $ICU$), are also available and therefore used to fit the model. We make the assumption that these variables $X_{k,obs}$ follow a discretized normal distribution with fixed standard deviation (Davies et al, 2021). In a first approximation, we assume that the mean equals the predicted values by the model $X_k$ (equations S2) times an observation rate $\rho_k$ (see Table S1) (Davies et al, 2021) and that the fixed standard deviation is proportional to its estimated highest value.

All variables are assumed to be independent conditional on the underlying epidemiological model, and therefore the model likelihood is the product of the likelihoods for all observed variables.

#### Inference

##### *Stochastic framework*

Equations (S1-S4) are considered in a stochastic framework solved with the Euler-Maruyama algorithm (Kloeden and Platen, 1999) implemented in the SSM platform (Dureau et al, 2013b).

##### *Estimation using the particle Markov Chain Monte Carlo (PMCMC) method*

The likelihood of our stochastic model being intractable, it was estimated with particle filtering method (Sequential Monte Carlo, SMC). With a given set of parameters, the SMC algorithm reconstructs sequentially the trajectory of the state variables and the time-varying parameters, and allows to compute the associated likelihood. Firstly, the distribution of the initial conditions of the system is approximated by a sample of particles. Then, at each iteration, the particles are projected according to the epidemiological and observation models up to the next observation point. To each of the particles a weight is associated, reflecting the quality of its prediction compared to the observation, and the total likelihood is updated. A resampling step using the weights is performed before the next iteration, in order to discard the trajectories associated with low weight particles.

In order to estimate the parameters of the system, the particle filter is embedded in a Markov Chain Monte Carlo framework, leading to the PMCMC algorithm (Andrieu et al, 2010). More precisely, the likelihood estimated by the SMC method is used within a Metropolis Hasting scheme (particle marginal Metropolis Hastings) (Andrieu et al, 2010). The proposal distribution is a Gaussian whose co-variance matrix is adapted following the framework described in Dureau et al (2013b).

The starting point of the MCMC chain is initialized using optimal values obtained from a simplex algorithm adapted for stochastic system (KSimplex), on a large number of parameter sets. Then, a pre-adaptation of the proposal co-variance matrix is performed with Kalman MCMC (KMCMC). The main idea behind these algorithmic improvements relies on less computationally costly algorithms in order to facilitate the exploration of parameter space. As our model is not Gaussian, we approximate the likelihood using the extended Kalman filter both in the simplex algorithm (KSimplex) (Dureau et al, 2013b) and in the MCMC (KMCMC) step (Dureau et al, 2013a). Then, the adaptive PMCMC is run on the output of the KMCMC step with 100,000 iterations and 2,000 particles for the estimates showed in figures included in the manuscript.

**PMCMC algorithm as implemented in SSM** (Dureau et al, 2013b)

In a model with $n$ observations and $J$ particles.
$q(.|\theta^{(i)})$ is the transition kernel

1. Initialize $\theta^{(0)}$
2. Using SMC algorithm, compute $\hat{p}(y_{1:n}|\theta^{(0)})$ and sample $x_{0:n}^{(0)}$ from $\hat{p}(x_{0:n}|y_{1:n}, \theta^{(0)})$
3. **for** $(i = 0: N-1)$ **do**
4. Sample $\theta^*$ from $q(.|\theta^{(i)})$
5. Using SMC algorithm, compute $L(\theta^*) = \hat{p}(y_{1:n}|\theta^*)$ and sample $x_{0:n}^*$ from $\hat{p}(x_{0:n}|y_{1:n}, \theta^*)$
6. Accept $\theta^*$ (and $x_{0:n}^*$) with probability $1 \wedge \frac{L(\theta^*)p(\theta^*)q(\theta^{(i)}|\theta^*)}{L(\theta^{(i)})p(\theta^{(i)})q(\theta^*|\theta^{(i)})}$
7. If accepted, $\theta^{(i+1)} = \theta^*$ and $x_{0:n}^{(i+1)} = x_{0:n}^*$. Otherwise, $\theta^{(i+1)} = \theta^{(i)}$ and $x_{0:n}^{(i+1)} = x_{0:n}^{(i)}$.
8. **end for**

| Parameters | Definitions | Prior or constant value | Prior or constant value | Prior or constant value |
|---|---|---|---|---|
| | | **Ile de France*** | **Ile de France**** | **Ireland** |
| $I_1(0)$ | Initial condition | U[10,1500] | U[10,1500] | U[5,100] |
| $S(0)$ | Initial condition | N=12278000 | N=12278000 | N=5176000 |
| $E_1(0), E_2(0), I_2(0), A_1(0), A_2(0)$ | Initial conditions | Use of steady-state conditions *** | Use of steady-state conditions *** | Use of steady-state conditions *** |
| $H_1(0), H_2(0), ICU(0), D(0), G(0), R(0)$ | Initials conditions | 0 | 0 | 0 |
| $\beta(0)$ | Initial condition of the transmission rate | 0.85 | 0.85 | 0.70 |
| $\nu$ | Volatility of the Brownian process | U[0.02,0.15] | U[0.02,0.15] | U[0.05,0.15] |
| $1/\sigma$ | average duration of the incubation period | tN[4,0.1,3,5] (Di Domenico et al, 2020) | tN[4,0.1,3,5] (Di Domenico et al, 2020) | tN[4,0.1,3,5] (Di Domenico et al, 2020) |
| $1/\gamma$ | average duration of the infectious period | tN[6,0.2,4.5,7.5] (eg Ferguson et al, 2020) | tN[6,0.2,4.5,7.5] (eg Ferguson et al, 2020) | tN[6,0.2,4.5,7.5] (eg Ferguson et al, 2020) |
| $1/\kappa$ | average hospitalization period | U[10,20] | U[10,20] | U[8,20] |
| $1/\delta$ | average time spent in ICU | U[10,20] | U[10,20] | U[8,20] |
| $\tau_A$ | fraction of asymptomatics | U[0.30,0.70] | U[0.30,0.70] | U[0.30,0.70] |
| $\tau_H$ | fraction of hospitalization | U[0.02,0.10] | U[0.02,0.10] | U[0.02,0.10] |
| $\tau_I$ | fraction of ICU admission | U[0.05,0.15] | U[0.05,0.15] | U[0.025,0.15] |
| $\tau_D$ | death rate | U[0.10,0.80] | U[0.10,0.80] | U[0.10,0.60] |
| $q_1$ | reduction in transmissibility | 1.5*$q_2$ but≤1 | 1.5*$q_2$ but≤1 | 1.5*$q_2$ but≤1 |
| $q_2$ | reduction in transmissibility | 0.55 (Li et al, 2020) | 0.55 (Li et al, 2020) | 0.55 (Li et al, 2020) |
| $q_I$ | reduction in ICU admission fraction | 0.05 | 0.05 | 0.10 |
| $q_D$ | reduction in death rate | 0.10 | 0.10 | 0.20 |
| $\rho_I$ | reporting rate for symptomatic infectious | U[0.01, 0.10] | U[0.01, 0.10] | U[0.02, 0.15] |
| $\rho_H$ | reporting rate for hospitalized people | U[0.95,1] | U[0.95,1] | U[0.95,1] |
| $\rho_{ICU}$ | reporting rate for ICU admission | 0.96 | 0.96 | 0.96 |
| $\rho_G$ | reporting rate for hospital discharge | 0.96 | 0.96 | 0.96 |
| $\rho_D$ | reporting rate for death | 0.98 | 0.98 | 0.98 |

*using hospital discharge data and **not using hospital discharge data
***steady-state conditions are defined by: $\frac{dE_1}{dt}=\frac{dE_2}{dt}=\frac{dI_1}{dt}=\frac{dI_2}{dt}=\frac{dA_1}{dt}=\frac{dA_2}{dt}=0$

**S1 Table.** (continued)

| Parameters | Prior or constant value | Prior or constant value | Prior or constant value | Prior or constant value |
|---|---|---|---|---|
| | **PACA** | **OC** | **NA** | **ARA** |
| $I_1(0)$ | U[10,500] | U[10,300] | U[10,200] | U[10,1000] |
| $S(0)$ | N=5055000 | N=5845000 | N=5957000 | N=5176000 |
| $E_1(0)$, $E_2(0)$, $I_2(0)$, $A_1(0)$, $A_2(0)$ | Use of steady-state conditions *** | Use of steady-state conditions *** | Use of steady-state conditions *** | Use of steady-state conditions *** |
| $H_1(0)$, $H_2(0)$, $ICU(0)$, $D(0)$, $G(0)$, $R(0)$ | 0 | 0 | 0 | 0 |
| $\beta(0)$ | 0.70 | 0.85 | 0.70 | 0.70 |
| $\nu$ | U[0.02,0.15] | U[0.02,0.15] | U[0.02,0.15] | U[0.02,0.15] |
| $1/\sigma$ | tN[4,0.1,3,5] (Di Domenico et al, 2020) | tN[4,0.1,3,5] (Di Domenico et al, 2020 | tN[4,0.1,3,5] (Di Domenico et al, 2020) | tN[4,0.1,3,5] (Di Domenico et al, 2020) |
| $1/\gamma$ | tN[6,0.2,4.5,7.5] (eg Ferguson et al, 2020) | tN[6,0.2,4.5,7.5] (eg Ferguson et al, 2020) | tN[6,0.2,4.5,7.5] (eg Ferguson et al, 2020) | tN[6,0.2,4.5,7.5] (eg Ferguson et al, 2020) |
| $1/\kappa$ | U[10,20] | U[8,18] | U[8,18] | U[10,20] |
| $1/\delta$ | U[10,20] | U[10,20] | U[14,24] | U[12,22] |
| $\tau_A$ | U[0.30,0.70] | U[0.30,0.70] | U[0.30,0.70] | U[0.30,0.70] |
| $\tau_H$ | U[0.02,0.15] | U[0.02,0.10] | U[0.02,0.10] | U[0.02,0.10] |
| $\tau_I$ | U[0.05,0.15] | U[0.05,0.20] | U[0.05,0.15] | U[0.05,0.15] |
| $\tau_D$ | U[0.10,0.60] | U[0.10,0.60] | U[0.10,0.60] | U[0.10,0.80] |
| $q_1$ | $1.5*q_2$ but≤1 | $1.5*q_2$ but≤1 | $1.5*q_2$ but≤1 | $1.5*q_2$ but≤1 |
| $q_2$ | 0.55 (Li et al, 2020) | 0.55 (Li et al, 2020) | 0.55 (Li et al, 2020) | 0.55 (Li et al, 2020) |
| $q_I$ | 0.05 | 0.05 | 0.05 | 0.05 |
| $q_D$ | 0.10 | 0.10 | 0.10 | 0.10 |
| $\rho_I$ | U[0.02, 0.15] | U[0.02, 0.15] | U[0.02, 0.15] | U[0.01, 0.10] |
| $\rho_H$ | U[0.95,1] | U[0.95,1] | U[0.95,1] | U[0.95,1] |
| $\rho_{ICU}$ | 0.96 | 0.96 | 0.96 | 0.96 |
| $\rho_G$ | 0.96 | 0.96 | 0.96 | 0.96 |
| $\rho_D$ | 0.98 | 0.98 | 0.98 | 0.98 |

**S2 Table.** Posteriors of the parameters for Ile-de-France region, Ireland, and four French regions: Provence Alpes Côte d'Azur (PACA), Occitanie (OC), Nouvelle-Aquitaine (NA), Auvergne Rhône Alpes (ARA).

| **Parameters** | **Definitions** | **Posterior** Median, [95%CI] | **Posterior** Median, [95%CI] | **Posterior** Median, [95%CI] |
|---|---|---|---|---|
| | | **Ile de France*** | **Ile de France**** | **Ireland** |
| $I_I(0)$ | Initial condition | 925, [443-1404] | 888, [487-1366] | 41, [18–80] |
| $\nu$ | Volatility of the Brownian process | 0.106, [0.075-0.143] | 0.106, [0.078-0.143] | 0.136, [0.108-0.149] |
| $1/\sigma$ | average duration of the incubation period | 3.99, [3.78-4.17] | 3.99, [3.81-4.21] | 3.99, [3.80-4.19] |
| $1/\gamma$ | average duration of the infectious period | 5.99, [5.63-6.34] | 6.12, [5.62-6.36] | 5.99, [5.63-6.38] |
| $1/\kappa$ | average hospitalizion period | 12.43, [10.65-14.48] | 14.98, [13.21-16.76] | 13.55, [12.05-15.17] |
| $1/\delta$ | average time spent in ICU | 15.86, [13.60-18.26] | 15.99, [13.74-18.55] | 17.48, [14.95-19.65] |
| $\tau_A$ | fraction of asymptomatics | 0.533, [0.325-0.692] | 0.537, [0.323-0.697] | 0.486, [0.308-0.688] |
| $\tau_H$ | average hospitalization period | 0.027, [0.021-0.043] | 0.023, [0.021-0.031] | 0.027, [0.020-0.046] |
| $\tau_I$ | fraction of ICU admission | 0.065, [0.055-0.100] | 0.076, [0.061-0.088] | 0.030, [0.023-0.045] |
| $\tau_D$ | death rate | 0.486, [0.425-0.525] | 0.504, [0.457-0.558] | 0.410, [0.362-0.462] |
| $\rho_I$ | reporting rate for symptomatic infectious | 0.023, [0.015-0.042] | 0.026, [0.016-0.035] | 0.092, [0.062-0.142] |
| $\rho_H$ | reporting rate for hospitalized people | 0.983, [0.955-0.998] | 0.982, [0.953-0.998] | 0.970, [0.951-0.998] |

*using hospital discharge data and **not using hospital discharge data

**S2 Table.** (continued)

| **Parameters** | **Posterior** Median, [95%CI] | **Posterior** Median, [95%CI] | **Posterior** Median, [95%CI] | **Posterior** Median, [95%CI] |
|---|---|---|---|---|
| | **PACA** | **OC** | **NA** | **ARA** |
| $I_I(0)$ | 138, [68-251] | 101, [49-231] | 91, [44-147] | 371, [171-729] |
| $\nu$ | 0.114, [0.092-0.143] | 0.123, [0.084-0.147] | 0.130, [0.105-0.148] | 0.131, [0.090-0.147] |
| $1/\sigma$ | 4.01, [3.82-4.20] | 4.01, [3.81-4.20] | 4.02, [3.81-4.20] | 4.00, [3.82-4.19] |
| $1/\gamma$ | 6.06, [5.70-6.38] | 6.00, [5.57-6.36] | 6.05, [5.63-6.50] | 5.98, [5.67-6.37] |
| $1/\kappa$ | 14.55, [13.17-16.04] | 11.07, [9.78-12.52] | 13.99, [11.95-16.72] | 14.95, [12.56-17.61] |
| $1/\delta$ | 13.54, [12.35-15.02] | 13.80, [11.83-15.44] | 16.51, [14.55-19.21] | 14.58, [12.25-17.45] |
| $\tau_A$ | 0.464, [0.314-0.664] | 0.530, [0.318-0.685] | 0.456, [0.323-0.678] | 0.495, [0.313-0.690] |
| $\tau_H$ | 0.023, [0.020-0.031] | 0.024, [0.021-0.036] | 0.024, [0.021-0.038] | 0.023, [0.021-0.036] |
| $\tau_I$ | 0.066, [0.058-0.080] | 0.076, [0.062-0.099] | 0.064, [0.052-0.089] | 0.060, [0.051-0.081] |
| $\tau_D$ | 0.412, [0.372-0.467] | 0.348, [0.305-0.396] | 0.434, [0.378-0.513] | 0.473, [0.411-0.542] |
| $\rho_I$ | 0.048, [0.034-0.071] | 0.026, [0.021-0.042] | 0.027, [0.022-0.042] | 0.023, [0.015-0.036] |
| $\rho_H$ | 0.988, [0.958-0.999] | 0.978, [0.952-0.998] | 0.980, [0.952-0.999] | 0.980, [0.952-0.998] |
